# Chinese moyamoya disease study:cohort profile

**DOI:** 10.1101/2023.03.20.23287508

**Authors:** Fang-Bin Hao, Cong Han, Gan Gao, Si-meng Liu, Min-jie Wang, Ri-Miao Yang, Zheng-Xing Zou, Dan Yu, Caihong Sun, Qian Zhang, Houdi Zhang, Qing-Bao Guo, Xiao-Peng Wang, Xuxuan Shen, Heguan Fu, JingJie Li, Zheng-shan Zhang, Xiang-Yang Bao, Jie Feng, Bin Li, Bin Ren, Hui Wang, Qian-Nan Wang, Songtao Pei, Bo Zhao, Zhenglong Zou, Yi-Qin Han, Huaiyu Tong, Zhenghui Sun, Miao Liu, Lian Duan

**Affiliations:** Chinese PLA General Hospital; 307 Hospital, PLA, The center for cerebral vascular disease, PLA; Medical School of Chinese PLA; (Department of Neurosurgery, Chinese PLA General Hospital (Former Department of Neurosurgery, the Fifth Medical Center of Chinese PLA General Hospital)); the Fifth Medical Center of Chinese PLA General Hospital; the Fifth Medical Centre, Chinese PLA General Hospital; the 307th Hospital of the Chinese People’s Liberation Army, The Fifth Medical Center of Chinese PLA General Hospital, Academy of Military Medical Science; 307 clinical college of Anhui Medical University); the Fifth Medical Center, Chinese PLA General Hospital; 307 Hospital, PLA; the 307th Hospital of the Chinese People’s Liberation Army,The Fifth Medical Center of Chinese PLA General Hospital; The 307th Hospital of the Chinese People’s Liberation Army, Academy of Military Medical Science; the Eighth Medical Center of Chinese PLA General Hospital; the Fifth Medical Centre of Chinese PLA General Hospital

**Author notes:** Corresponding author: Lian Duan, Miao Liu, Zhenghui Sun. These authors have contributed equally to this work and share first authorship (Fang-Bin Hao, Cong Han, Gan Gao).

## Abstract

**Background:** The etiology and natural course of moyamoya disease (MMD) remain unknown. This study evaluated the natural course and etiology of MMD using data from the Chinese MMD (C-MMD) cohort study.

**Methods:** The C-MMD cohort consisted of 6,167 patients with MMD treated at our center over the past 20 years. We analyzed the medical history and laboratory and imaging examination results of the patients at different stages of the disease to identify common features of MMD.

**Results:** The median age for symptom onset was 32 years. The age distribution was bimodal; the highest peak was between ages 35 and 45 years, with a smaller peak between ages 3 and 9 years. The female-to-male ratio was 1:1. The disease occurred mainly in the Han people and was rarely observed in minority nationalities. In our cohort, a transient ischemic attack was the most common initial clinical manifestation (46.3%); others included infarction (25.0%), hemorrhage (15.1%), and headache (7.9%).

**Conclusions:** The C-MMD cohort is currently one of the largest single-center MMD cohorts in the world. This study provides baseline data for future research on the etiology and natural course of MMD.

**Clinical Trial Registration:** This study has been registered in the Chinese Clinical trial registry (registration number: ChiCTR2200064160).

## 1 Introduction

Moyamoya disease (MMD) is a primary chronic cerebrovascular disorder characterized by gradual stenosis and occlusion of the bilateral distal internal carotid arteries and the proximal portion of the anterior or middle cerebral arteries.^1,2^ MMD is the most common pediatric cerebrovascular disease and the leading cause of stroke in children and young adults in Japan, Korea, and China.^3-5^ Furthermore, MMD can manifest as ischemic or hemorrhagic strokes, seizures, and cognitive impairment, resulting in a high prevalence of disability and death.^6^ The high disability rate of MMD also burdens the patients’ families and society.

MMD is more prevalent in East Asian populations, such as Japan, Korea, and China, than in Europe, America, and Africa.^4,7-12^ Furthermore, the incidence of MMD has been increasing for the last few years owing to increased numbers of evaluations and thus disease recognition. For example, the MMD incidence rate in Japan was 1.13/100,000 people in 2012, showing a nearly two-fold increase from 2003.^5,13^ This trend has also been observed in Korea, with an incidence rate of 18.1/100,000 people in 2013 versus 4.3/100,000 in 2005, a nearly three-fold increase.^4^ China has the largest population in the world, and recent epidemiological data reported an annual increase in MMD from 2016 to 2018, from 0.88 to 1.47/100,00 people.^7^ The MMD incidence has also increased in the United States from 1988 to 2004; 2,247 patients with MMD were admitted to the hospital, but 7,473 patients were hospitalized from 2005 to 2008. Thus, the incidence rate increased from 0.086/100,000 people in 2005 to 0.57/100,000 in 2012.^14-17^

MMD is a serious disease with a poor prognosis; thus, the social burden increases with the number of MMD cases. However, the etiology, pathogenesis, and clinical course of MMD are unclear.^18^ Consequently, suitable animal models have not been established, and basic research has not progressed beyond the molecular and cellular levels.^19^ MMD susceptibility genes have been identified, such as ring finger protein 213-coding gene *RNF213*; however, no apparent causative etiology factor has been found.^20,15,16^ Moreover, the clinical manifestations of MMD vary greatly, with considerable differences in the onset time, progression rate, disease severity, and therapeutic effect among patients. In addition, the genetic background of patients with MMD in China is complex; therefore, MMD subtypes may exist.^7,21^ Finally, few large-scale studies have been conducted on MMD. Thus, little is known about the clinical course.

Clarifying the natural course of the disease and its etiology is an urgent issue. Over the last 20 years, our institution has collected clinical information on patients with MMD treated at our center, establishing the Chinese MMD (C-MMD) cohort. Therefore, this study assessed the C-MMD cohort to elucidate the clinical course and the etiology of MMD.

## 2 Methods

### 2.1 Study population

The C-MMD cohort study is a Chinese nationwide observational study comprising patients registered at PLA General Hospital and then followed up in a real-world setting. The inclusion criteria were based on the 2021 Guidelines for the diagnosis of MMD.^22^

The inclusion criteria were: (1) The presence of stenosis or occlusion of the intracranial internal carotid artery or proximal portions of the anterior or middle cerebral arteries on digital subtraction angiography (DSA) or magnetic resonance angiography (MRA); (2) observation of abnormal vascular networks during the arterial phase of the disease near occlusive or stenotic lesions on DSA or abnormal vascular networks in the basal ganglia on MRA; and (3) bilaterality of findings (1) and (2).

The exclusion criteria were (1) incomplete medical records and (2) moyamoya syndrome caused by atherosclerosis, autoimmune diseases, meningitis, brain tumor, Down syndrome, neurofibromatosis, head trauma, cerebrovascular injury due to head radiation irradiation, and other clearly defined causes.

### 2.2 Data collection

Clinical data and biological samples were collected. Clinical data included patients’ medical history, imaging examination, laboratory results, and cognitive function results. Table 1 presents the specific assessment contents.

#### 2.2.1 Clinical data

##### 2.2.1.1 Medical history data

We collected demographic and clinical data, including age, sex, place of birth, time of admission, marital status, occupation, first symptoms, co-existing diseases, past medical history, familial history, cerebrovascular events, surgery, and drug therapy. The first symptom type, onset time, and attack frequency were also recorded. Cerebrovascular events were determined based on the clinical history and an imaging examination. Modified Rankin Scale and National Institute of Health Stroke Scale scores were used for evaluating survival status and scored by professionally trained neurologists.

##### 2.2.1.2 Imaging data

A dedicated individual gathered C-MMD imaging data systematically starting in 2002. All images were stored in a dedicated radiology film library and backed up electronically. The C-MMD study collected patients’ DSA, magnetic resonance imaging, MRA, perfusion-weighted imaging, contrast-enhanced high-resolution magnetic wall imaging, computerized tomography, emission computerized tomography, positron emission tomography-computed tomography, and transcranial Doppler ultrasound images. The DSA Suzuki grade,^23^ post-operative Matsushima grade,^24^ and MMD collateral grading score^25^ were also analyzed.

##### 2.2.1.3 Cognitive function

Systematic cognitive function assessments were conducted. Cognitive data were collected by a qualified and trained specialist (Hou-Di Zhang). Nine cognitive assessment scales were used, including the Montreal Cognitive Assessment,^26,27^ the Mini-Mental State Examination,^28^ Activity of Daily Living Scale,^29,30^ the 17-item Hamilton Rating Scale for Depression,^31^ clock drawing test,^32^ the Chinese auditory verbal learning test,^28,33^ Trail Making Test A and B,^34^ and Boston Naming Test.^35^

##### 2.2.1.4 Laboratory examination

Patients were routinely tested on the second day of each admission for routine blood tests, blood biochemical tests, and immune tests.

#### 2.2.2 Biological sample collection

Biological samples were retained for further investigation to assess the etiology of MMD and identify biomarkers that reflect disease progression. A written informed consent form approved by the Ethics Committee of PLA General Hospital was signed by the patient or their guardian before biological sample collection. Blood samples (4 mL EDTA tube and 4 mL SST tube) and urine samples (10 mL) were collected and stored in four parts in a -80 °C freezer.

### 2.3 Follow up

The C-MMD study design was a historical prospective cohort with follow-up from the time of first admission to the hospital. The follow-up methods included in-hospital and out-of-hospital follow-ups. In-hospital follow-ups included in-hospital and out-patient follow-ups. Out-of-hospital follow-ups consisted of email, telephone, WeChat, and electronic medical record follow-ups. Follow-ups were mainly conducted by one trained researcher (Qian Zhang) and performed three and six months after discharge and every year after that.

### 2.4 Data management and quality control

A web-based electronic data capture system was used to capture all clinical data. An independent data safety and monitoring committee was established to supervise the conduct of the C-MMD study, composed of clinicians and statistical experts. Regular meetings occurred to ensure that the study design and data statistics were scientific. The cohort was quality controlled by the professional statistics team of the Department of Statistics and Epidemiology, Graduate School, Chinese PLA General Hospital, and all data were entered by professionally trained medical staff and epidemiology professionals. All information was checked by the C-MMD data safety and monitoring committee.

The Ethics Committee of the General Hospital of the Chinese People’s Liberation Army approved this study (No. KY-2022-9-69-1). This study has been registered in the Chinese Clinical trial registry (registration number: ChiCTR2200064160).

## 3 Results

### 3.1 Demographic data

The C-MMD cohort study enrolled 6,167 patients with MMD. All patients were admitted to the Department of Neurosurgery at the Chinese PLA General Hospital between April 2002 and May 2022. Each patient had a unique patient ID number and complete clinical records available in the hospital’s database. The patients were from 32 provinces in China. The primary ethnic group was Han Chinese (98.9%); Mongolian, Manchu, and 11 other minority nationalities accounted for 1.1%. The male-to-female ratio was nearly 1:1 (3,096:3,068). The youngest patient was three months old at the onset time, and the oldest was 75 years old. The mean age at admission was 34.78 years, and the age distribution was bimodal (Figure 1). More women were aged 15 to 24 than men (female-to-male ratio, 3:2 [294:189]). The age distributions of other groups were similar (Figure 2).

**Figure 1.**
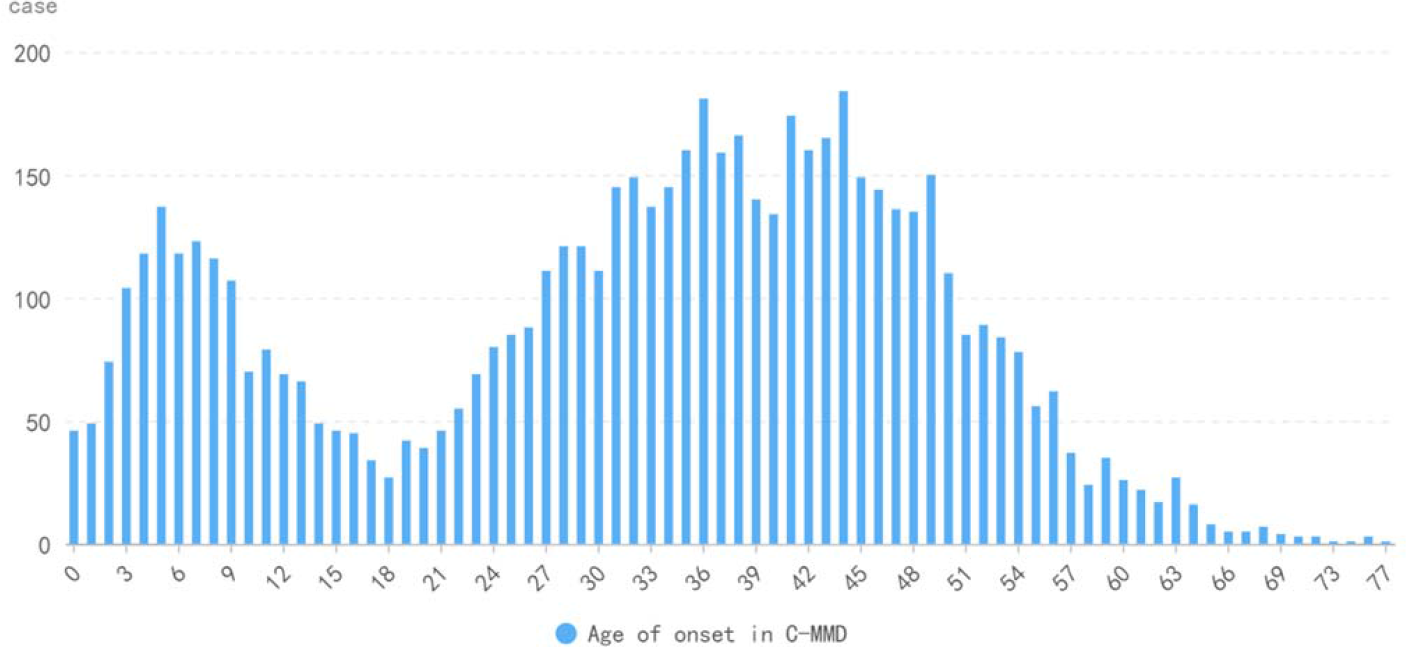
Age distribution of C-MMD patients. C-MMD: Chinese moyamoya disease

**Figure 2.**
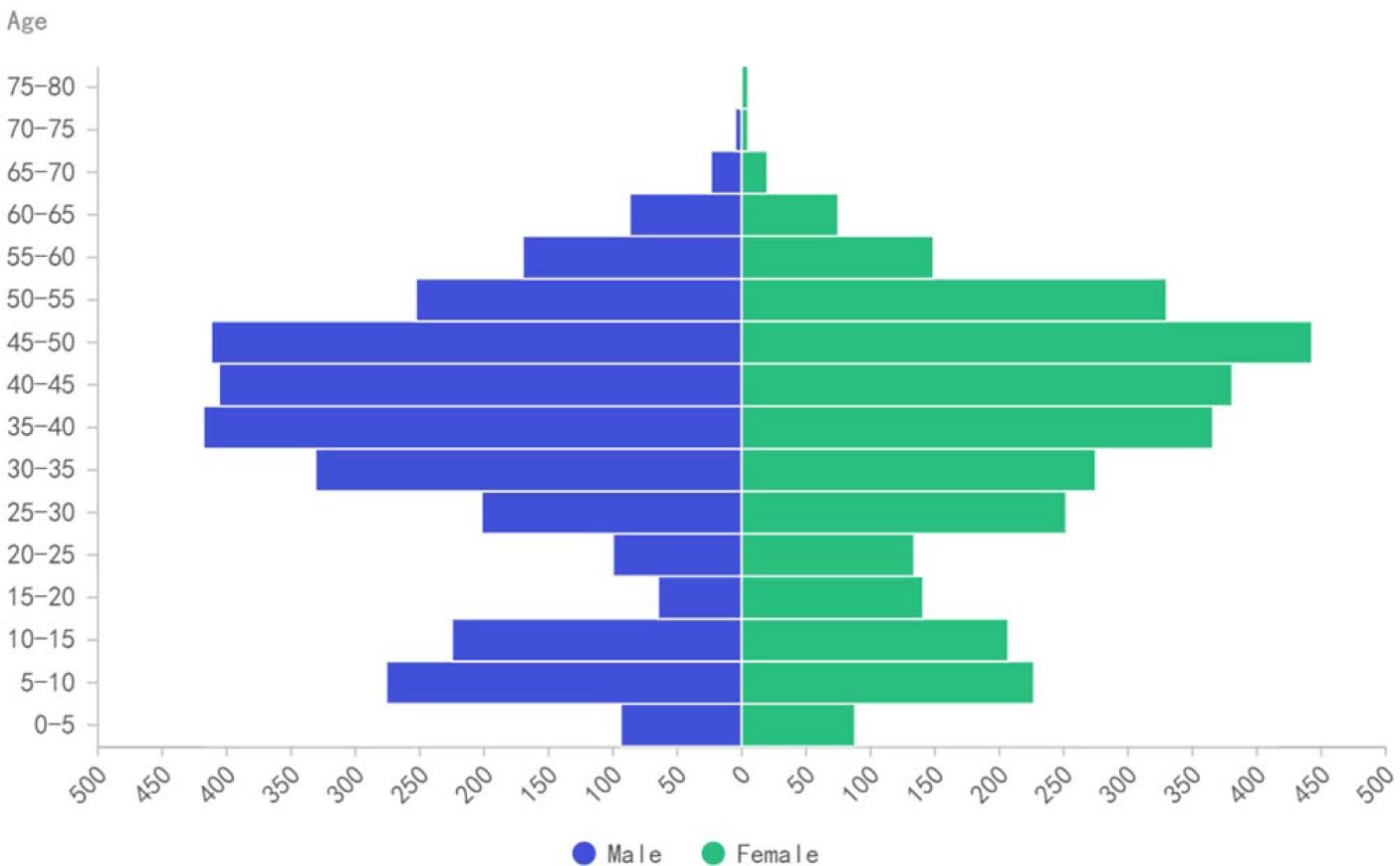
Distribution of male-to-female ratios in C-MMD patients at different ages. C-MMD: Chinese moyamoya disease

The primary onset symptom was a transient ischemic attack (TIA). The proportion of patients with TIA was comparable to that of Japanese and South Korean patients but higher than that of European and American populations. Compared to the national hospital MMD survey led by Tiantan Hospital, the proportion of patients with cerebral infarction and cerebral hemorrhage was slightly lower than those with TIA but similar to that of the Japanese epidemiological survey (Figure 3). Most patients in the C-MMD cohort were from the Hebei, Shandong, and Henan provinces.

**Figure 3.**
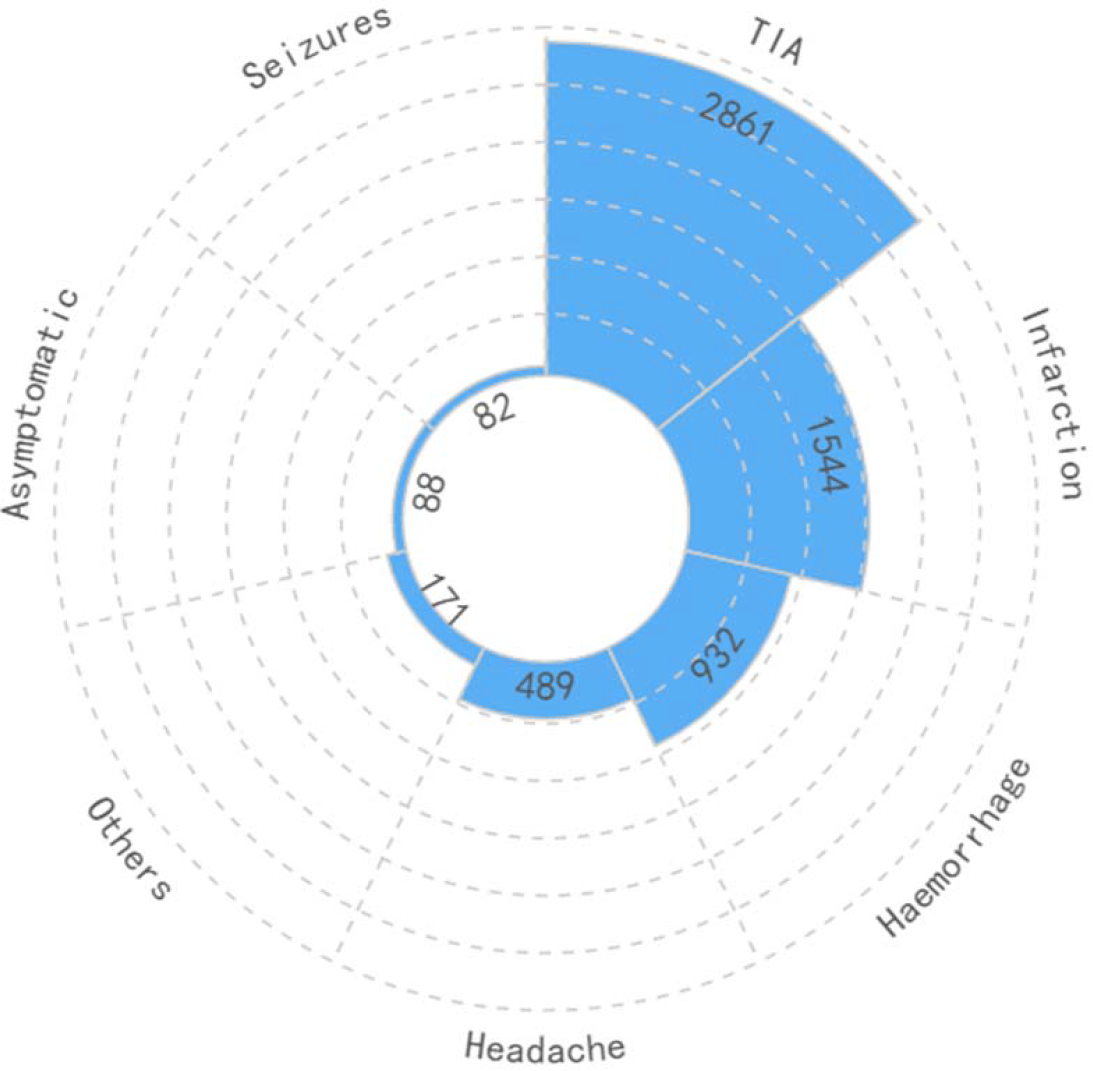
Distribution of first symptoms in C-MMD patients. C-MMD: Chinese moyamoya disease; TIA: transient ischemic attack

### 3.2 Familial occurrence

The proportion of patients with a family history was 7.1% (442/6,167). Of the 6,167 patients with MMD, 122 patients had siblings with an MMD diagnosis, 68 had mothers with MMD, and 102 had fathers with MMD. Cerebral or MRA exams were performed on all family members mentioned earlier.

### 3.3 Follow-up

Overall, 61.6% (3801/6167) of the patients underwent in-hospital follow-ups. The rate of overall follow-up using standardized and active out-of-hospital follow-up assessments was 92.6%. The primary follow-up concerns were the changes in symptoms, laboratory results, imaging indicators, and drug use in other hospitals.

## 4 Discussion

MMD was first reported in 1957^2^ and is highly prevalent among people of Asian descent, especially those of East Asia. Furthermore, as MRA and computed tomography angiography examinations have become more accessible, more cases of MMD have been discovered. For instance, in Japan, approximately 0.07% of individuals in the general population have MMD, with an estimated prevalence of 50.7/100,000 people.^36^ Korea’s Ministry of Health and Welfare reported a 15% increase in the prevalence of MMD from 2004 to 2008 (from 5.2/100,000 people to 9.1/100,000), reaching 18.1/100,000 in 2013.^4^ Additionally, recent epidemiological studies in China reported that the number of new MMD cases increased from 0.88/100,000 people in 2016 to 1.47/100,000 in 2018.^7^ European and American countries experience lower rates of MMD than East Asian countries, but their incidence rates have also increased over the past few years.^15^ For example, the U.S. Department of Health Insurance and the National Inpatient System reported 2,247 MMD-related hospitalizations from 1988 to 2004 and 7,473 hospitalizations from 2005 to 2008.^14,16^

Japanese and Korean studies have reported two onset age peaks for MMD: first in childhood (average age: 5 to 10 years), then in adulthood (average age: 40 to 50 years).^4,5^ In the C-MMD cohort, the onset age varied from 0 to 77 years, with two peaks similar to those reported in the Japanese and South Korean studies. Conversely, an epidemiological MMD study led by Tiantan Hospital that included 47,443 patients admitted to 1,312 hospitals in China between 2016 and 2018^7^ reported a unimodal age distribution. We propose two explanations for these conflicting results. First, there are no standard MMD symptoms; thus, some children with mild early symptoms may not have been diagnosed and treated at that time. Second, our center treated more children with MMD because we began systematically treating patients with MMD earlier and gave great attention to detecting and treating children with MMD.

Asian epidemiological surveys from Japan reported more female patients with MMD than men (female-to-male ratio, from 1.8:1 to 2.2:1),^5,13^ as did studies of heterogeneous American populations (2.5:1 and 1.8:1)^14^ and a white European population (4.25:1).^37^ The C-MMD cohort ratio was 1:1, consistent with Chinese epidemiology research.^7^ Hence, Chinese patients with MMD may have a unique genetic background.

Patients with MMD can present with various symptoms, mainly classified as cerebral ischemia, cerebral hemorrhages, and others.^38,39^ However, improved screening methods have increased the number of patients with MMD without obvious clinical symptoms.^36^

The molecular etiology and natural progression of MMD and the lack of effective animal models necessitate a large cohort study to summarize clinical characteristics and explore possible pathogenic mechanisms.^6^ As a result, our team collected clinical and biological data on 6,167 patients with MMD. The C-MMD study is an ongoing nationwide real-world observational study registered in PLA General Hospital in China. Consequently, the C-MMD study is one of the most extensive MMD cohort studies with some notable strengths. First, this study collected various forms of information, including patients’ symptomatology, imaging, laboratory examination, and cognitive function data, resulting in a compressive collection of data that enables disease severity and survival status evaluations from multiple perspectives. Second, the follow-up time was extensive, with a low loss-to-follow-up rate. This study systematically includes patients with MMD starting in 2002, and over the past 20 years, our team has maintained good doctor-patient relationships. Furthermore, many patients were hospitalized for multiple follow-up visits, providing good conditions for monitoring the natural course of MMD. Third, we built a valuable biobank and subsequently identified some MMD susceptibility genes in the Chinese population through large sample genome-wide association studies. At present, 3,952 biological samples have been collected for the C-MMD study, providing good scope for further studies on the etiology of and biomarkers for MMD.

## 5 Limitations

This study also has several limitations. First, the C-MMD study was conducted in Beijing, the capital of China; thus, many patients were referred to our hospital. Therefore, some medical records regarding the first onset were missing. Second, this single-center historically prospective hospital-based study may have selection and information bias.

## 6 Conclusion

C-MMD is currently one of the largest single-center MMD cohorts in the world. This study provides baseline data for future research on the etiology and natural course of the disease.

## Data Availability

C-MMD cohort members welcome and encourage collaboration to maximize the use of data and samples. The data is currently not freely available in the public domain owing to sensitive information in the study database. However, specific proposals for possible collaborations are welcome. Researchers interested in collaboration and further information are invited to contact the corresponding author by email at: (Duan lian).

## Non-standard Abbreviations and Acronyms

C-MMD: Chinese moyamoya disease
DSA: Digital subtraction angiography
MRA: Magnetic resonance angiography
MMD: Moyamoya disease
TIA: Transient ischemic attack

## Acknowledgments

We would like to extend our sincere gratitude to all participants in the C-MMD study, as well as all members of the research team. We would like to thank Editage (www.editage.cn) for English language editing.

## Author contributions

The C-MMD cohort is established and the data collection is conducted by the steering committee with teams (F.H., C.H., G.G., S.L., M.W., R.Y., Z.Z., D.Y., Q.Z., H.Z., Q.G., X.W., X.S., H.F., J.L., Z.Z., X.B., J.F., B.L., B.R., H.W., Q.W., S.P., B.Z., Z. Z., Y.H., H.T., Z.S., M.L. and L.D.). L.D., M.L., C.H., and F.H. designed this study. F.H. supervised the research and drafted the manuscript. M.L. and F.H. organized and analyzed the data. All authors contributed to data analysis, assisted with data interpretation, and critically revised and approved the manuscript.

